# Frequency of preclinical Alzheimer’s disease in Japanese clinical study settings: a meta-analysis

**DOI:** 10.1101/2022.03.22.22272449

**Authors:** Kenichiro Sato, Yoshiki Niimi, Ryoko Ihara, Kazushi Suzuki, Atsushi Iwata, Takeshi Iwatsubo

**Affiliations:** Department of Neuropathology, Graduate School of Medicine, The University of Tokyo; Unit for Early and Exploratory Clinical Development, The University of Tokyo Hospital; Department of Neurology, Tokyo Metropolitan Geriatric Medical Center Hospital; Division of Neurology, Internal Medicine, National Defense Medical College

**Keywords:** preclinical Alzheimer’s disease, prevalence, meta-analysis, Asian cohort

## Abstract

**Background:** Many of earlier studies reporting the frequency of preclinical Alzheimer’s disease (AD) among cognitive normal (CN) individuals have based on the clinical study cohort from Western countries, and it has not been validated whether the frequency of preclinical AD in Asian country cohort may differ from that in Western countries.

**Methods:** We conducted meta-analysis of earlier literature and original data of 4 Japanese cohort study (i.e., AMED-Preclinical, Brain/Minds Aging Imaging Study, J-ADNI, and A4 study screening conducted in Japan), among which we incorporated cognitive normal (CDR = 0) clinical study participants who were examined their amyloid status by amyloid-PET or CSF.

**Results:** In total, among the reviewed 658 Japanese CN participants from 10 different groups, 103 turned out to be amyloid positive, and the estimated overall frequency of preclinical AD (of any stage) was 17.2% (95% CI: 12.0 – 23.0%) with a high statistical heterogeneity of I^2^ = 64%. In meta-regression, being the Japanese cohort was barely significantly associated with the lower frequency of preclinical AD (*p = 0.048*).

**Conclusions:** Our study suggested that currently there is no robust evidence to support the lower frequency of preclinical AD in clinical study settings of Japan than that of Western countries.

## Background

Preclinical Alzheimer’s disease (AD), which corresponds to positive brain amyloid beta (Aβ) accumulation in healthy individuals without an evidence of cognitive decline^1-3^, is getting focused as the target of clinical trials aiming to develop disease-modifying therapies for AD^4^.

Relatively low rate of positive Aβ among cognitive normal elderly individuals (e.g., approximately one-third^3, 5^) is one of the major barriers in recruiting such preclinical AD participants to the clinical trials. The frequency of the “preclinical AD” was reported as 38% in NA-ADNI cohort study^3^ or 33% in AIBL cohort study^6^, and 22% by a recent meta-analysis^7^. However, many of the earlier studies reporting the frequency of preclinical AD have based on the clinical study cohort from Western countries^5, 7^, and the frequency in Asian country cohorts remain much of uncertainties.

In Japan, J-ADNI study, a large prospective observational study conducted in Japan^8^, had reported a frequency of 22.6% of preclinical AD at any stage^9^, which was lower than that of similar large-scale cohort studies such as the above NA-ADNI or AIBL. As the frequency of preclinical AD in clinical study cohort participants should vary depending on the participants’ age distribution and the frequency of having *APOE*-e4 allele(s)^5, 9^, this difference in the frequency may be due to the coincidental variance in the included participants. Meanwhile, it is reported that the estimated prevalence of *APOE*-e4 in Asia (including Japan) is lower than that in North America or Central/Northern Europe^10^, so that the prevalence of preclinical AD in general population may also be lower in Japan than in Western countries, when compared between the cognitive normal elderly individuals of the same range of age.

Participants in clinical study cohort are selected from general population based on the predefined inclusion/exclusion criteria, so that the frequency in the clinical study cohort may not be such a robust epidemiological measure unlike prevalence in general population. However, accurately estimating the frequency of preclinical AD in clinical study settings is still important because clinical trials for AD therapy are indeed conducted based on such study cohort. If the frequency of preclinical AD in Japanese cohort turned out to be significantly lower than that of Western country cohorts, whichever the reasons for the difference are, we need to recruit significantly larger number of participants in order to confirm the similar level of treatment efficacy as of the Western cohorts.

Alongside the above J-ADNI study, there are some additional Japanese cohort studies screening for the preclinical AD individuals being conducted but have not been not published yet. By meta-analyzing these Japanese cohort studies in addition to earlier cohort studies, in this study, we aim to evaluate whether the frequency of preclinical AD in the clinical study settings in Japan is different from Western countries, using meta-regression analysis.

## Methods

### Reviewing process: overview

In this study, we reviewed two types of data source in order to extract cohort summary to use in the meta-analysis: original data of several preclinical AD clinical studies (Figure 1A), and earlier literatures reporting the frequency of preclinical AD (Figure 1B). Based on the NIA-AA criteria^1, 11^, preclinical AD considered in this meta-analysis was defined as those with positive evidence of Aβ accumulation in brain among the cognitive normal - of which definition includes the Clinical Dementia Rating (CDR) = 0^12^. Since the degree of subtle cognitive decline varies depending on the additional inclusion criteria of each cohort (e.g., some studies require MMSE scores 27-30 for eligibility, while others not), and some additional biomarker tests to classify preclinical AD staging^1^ 1-3 (e.g., structural brain MRI, CSF p-tau or t-tau) are not always reported, in this study we meta-analyzed about the frequency of preclinical AD across all stages 1-3. Each cohort was classified depending on whether it is from non-Asian countries or Japan, and if one study is comprised of multi-national cohorts (e.g., solanezumab in A4 study^13, 14^ was administered for United States cohort and Japanese cohort), we separately treated each cohort depending on the country of origin. The included non-Asian cohorts were, actually as a result, all from Western countries.

**Figure 1.**
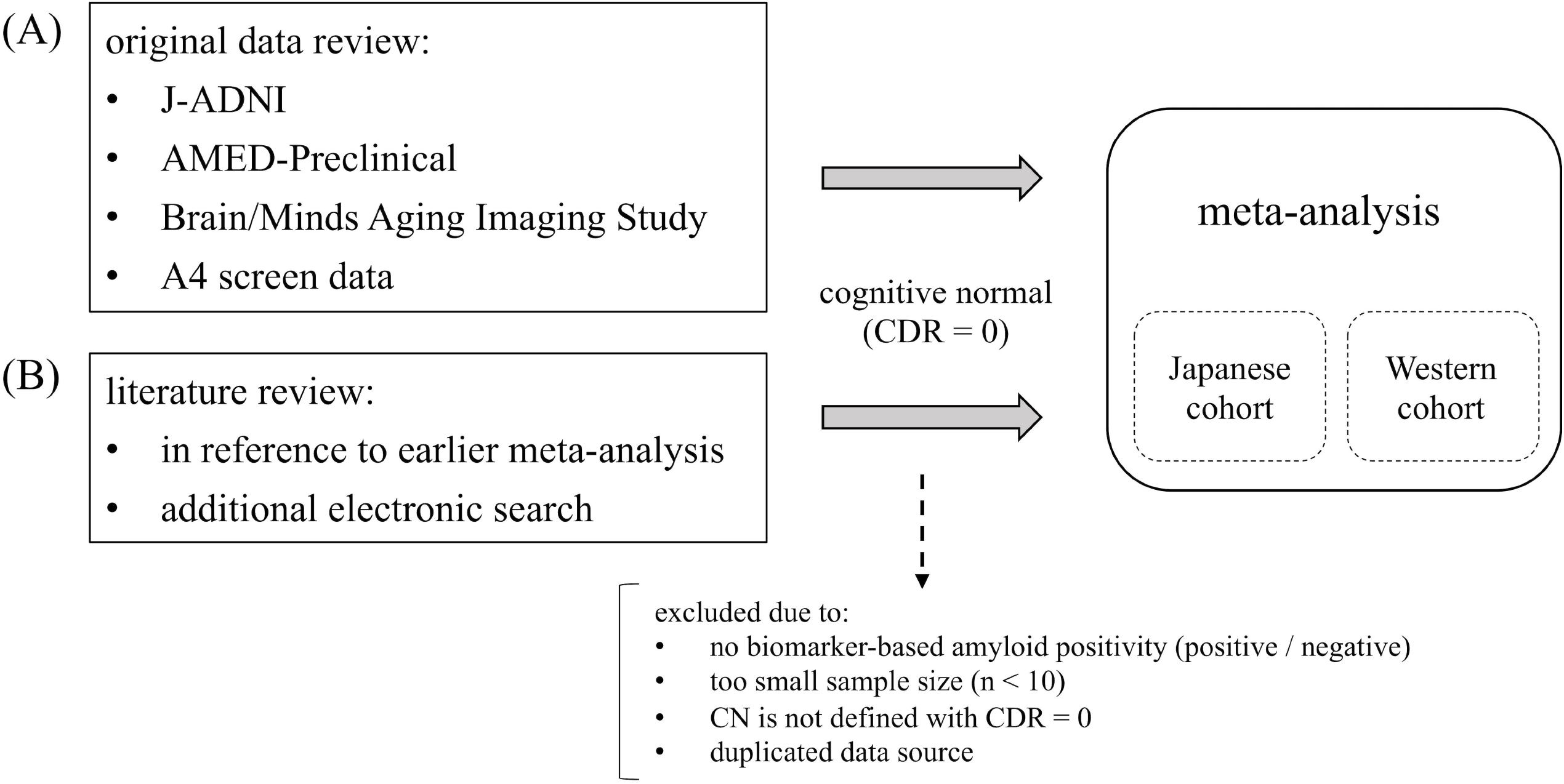
Reviewing process. We reviewed two types of data source in order to extract cohort summary to use in the meta-analysis: (A) original data of several preclinical AD clinical studies, and (B) earlier literatures reporting the frequency of preclinical AD. Abbreviations: AD, Alzheimer’s disease; CDR, Clinical Dementia Scale; CN, cognitive normal

### Review of original data

For reviewing the original data, we included 4 different cohorts (Figure 1A) which were conducted in Japan in order to detect and investigate the preclinical AD among cognitive normal individuals, as follows: 1) Japanese Alzheimer’s disease Neuroimaging Initiatives (‘J-ADNI’)^8^, 2) Clinical and neuroimaging study on preclinical Alzheimer’s disease (‘AMED-Preclinical’), 3) Brain Mapping by Integrated Neurotechnologies for Disease Studies: Human Brain Aging Imaging Study (‘Brain/Minds Aging Imaging Study’), and 4) A4 study screening^13, 14^.

All these cohorts originally define the similar inclusion criteria, where cognitive normal (CN) participants must be aged 60-, both sex, their CDR-global = 0, and must not have significant neuropsychiatric findings or the significant past history of neuropsychiatric diseases. We reviewed data of participants who were determined as CN at the time of original study participation, and included the CN participants who had received either amyloid PET or CSF Aβ_42_ examinations. We retrieved the summary of amyloid status (positive rate among all tests), mean age, sex proportion, and the mean MMSE score (if applicable) from each cohort group.

The J-ADNI was launched in 2007 as a public-private partnership, led by Principal Investigator Takeshi Iwatsubo, MD. The primary goal of J-ADNI was to test whether clinical and neuropsychological assessment, together with biological markers including, but not limited to, serial magnetic resonance imaging (MRI) and positron emission tomography (PET), could be combined to measure the progression of MCI and mild AD in the Japanese population. The original data of J-ADNI study is available from National Bioscience Database Center (NBDC), with approval from its data access committee (https://humandbs.biosciencedbc.jp/en/hum0043-v1). The inclusion/exclusion criteria used in J-ADNI largely correspond to those for NA-ADNI^15^ and they are described at: https://upload.umin.ac.jp/cgi-open-bin/ctr_e/ctr_view.cgi?recptno=R000001668. Details are described in our previous reports^8, 9, 16-18^. Since many of the CN participants in J-ADNI cohort did not receive both PiB-PET and CSF, so we made two subgroups depending on the modality of amyloid tests (PET-based or CSF-based). For interpreting the PiB-PET result, “positive” or “equivocal” results were determined as positive Aβ accumulation in brain, according to the earlier report^9^. For interpreting CSF Aβ result, CSF Aβ_42_ < 333 pg/mL was determined as positive Aβ accumulation in brain^9^.

The AMED-Preclinical is a multi-center observational study launched on 2016 aiming to discriminate MCI individuals at risk for development of Alzheimer dementia, as well as preclinical AD without clinical manifestations. The summary results of the AMED-Preclinical have not been published yet, but the original data is publicly distributed at: https://humandbs.biosciencedbc.jp/en/hum0235-v1. The inclusion/exclusion criteria used in the AMED-Preclinical is mostly correspond to those for J-ADNI as described above, and they can be confirmed at: https://upload.umin.ac.jp/cgi-open-bin/ctr_e/ctr_view.cgi?recptno=R000022596.

The Brain/Minds Aging Imaging Study is another observational study launched on 2015 aiming to apply functional MRI methodologies to the very early stage of Alzheimer disease, and to acquire MRI imaging data including structural MRI, resting state functional MRI, diffusion tensor imaging and arterial spin labeling in MCI or preclinical AD individuals. The summary results of the Brain/Minds Aging Imaging Study have not been published yet, but the original data is publicly distributed at: https://humandbs.biosciencedbc.jp/en/hum0250-v1. The inclusion/exclusion criteria used in the Brain/Minds Aging Imaging Study is less stringent than that in the J-ADNI and AMED-Preclinical in terms of its eligible age (60-90 years old) and the cognitive inclusion criteria (e.g., MMSE or WMS-R are not required to be in specific range for eligibility). They can be confirmed at: https://upload.umin.ac.jp/cgi-open-bin/ctr_e/ctr_view.cgi?recptno=R000022596.

The A4 screening data is the result of amyloid-PET screening process in A4 trial, a phase-3 RCT of solanezumab for preclinical AD individuals^13^, and we downloaded it from the LONI website (https://ida.loni.usc.edu/login.jsp) under permission. We used the same eligibility as of an earlier report^14^, as follows: 65-85 aged CN (CDR-global = 0) individuals with MMSE 25-30 and logical memory delayed recall (IIa) 6-18, and the amyloid positivity was determined by the standardized uptake value ratio (SUVr)^19^ ≥ 1.15. Excluding Asian cases, we used the rest of A4 screening cases as one of the Western cohorts to include into the meta-analysis.

### Review of earlier literatures

Next, we reviewed earlier literatures published since 2008 to 2020 to compare the frequency of preclinical AD between Japanese cohorts and Western cohorts by meta-regression analysis. Since similar systematic review / meta-analysis study regarding the frequency of preclinical AD in 2008-2018 has already been conducted elsewhere^5, 7^, we performed additional hand search for PubMed (https://pubmed.ncbi.nlm.nih.gov) on early December 2020 to complement uncovered literatures from the previous systematic review reports. The search term in PubMed was as follows: (“preclinical” OR “asymptomatic” OR “cognitively normal”) AND “Alzheimer*” AND (“biomarker” OR “abeta” OR “amyloid-beta” OR “amyloid”) AND (“PET” OR “positron emission tomography” OR “CSF” OR “cerebrospinal fluid” OR “amyloid PET”) AND (2008:2020[pdat]). Thus, this study is not a systematic review, so that we have not registered our study to PROSPERO (https://www.crd.york.ac.uk/prospero/).

The eligibility criterion of earlier literatures to include into the current study were defined as follows: 1) studies which report the amyloid test results in binary (positive /or negative) for cognitive normal (CN) individuals, 2) studies whose CN criteria includes CDR-global = 0, 3) studies of which sample size is larger than 10, and 4) studies whose data source are not duplicated from others. We included the results from observational cohort study or from the clinical trial screening for amyloid tests. All the included literatures are summarized in Table 1. We retrieved the summary of amyloid status (positive among all tests), mean age, sex, and mean MMSE score from each literature when applicable.

**Table 1.**
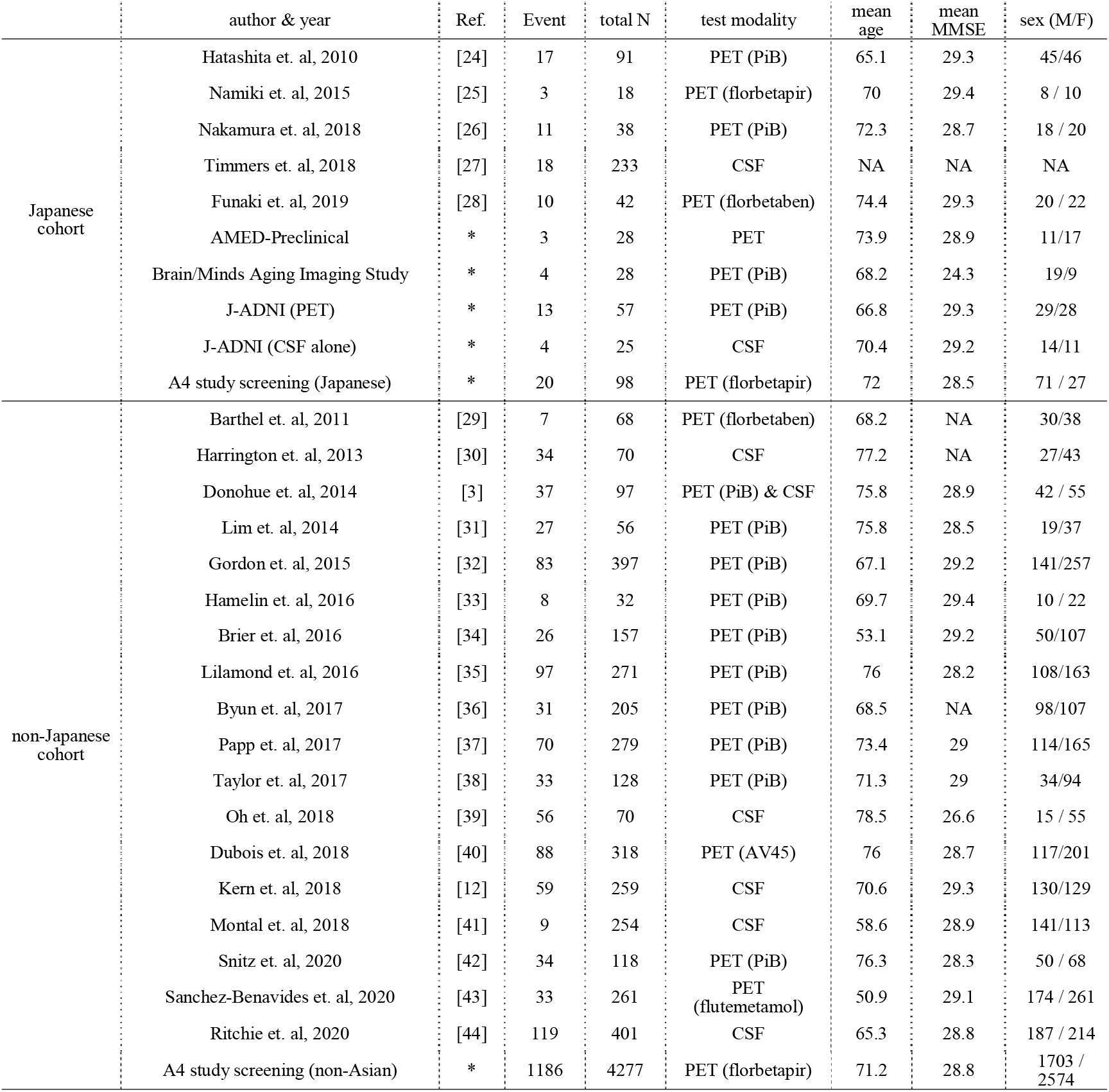
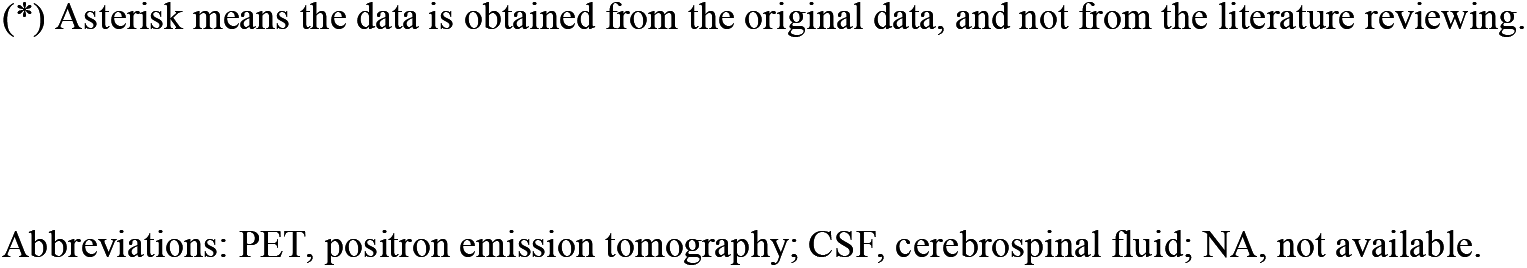
Summary of the reviewed earlier literatures and the original data

### Statistical analysis

All the data handling and analyses were performed using R 3.5.1 (R Foundation for Statistical Computing, Vienna, Austria). A p-value level < 0.05 was considered as statistically significant if not mentioned otherwise. We used R package *meta*^20^ and *metafor*^21^ for the meta-analysis of single proportion of preclinical AD among each cohort. Random-effect model was used, and the statistical heterogeneity of the included data was assessed by I^2^ (%) measure. Publication bias was assessed by Egger’s test^22^. Meta-regression was conducted to assess whether the frequency of preclinical AD in Japanese cohorts is significantly different from that in non-Japanese (i.e., Western country) cohorts, while adjusting mean age of cohorts. For drawing spline in the plot, we used R package *mgcv*^23^.

### Ethics

Reviewing the data included in this study has been approved by the University of Tokyo Graduate School of Medicine institutional ethics committee (ID: 11628-(3)). Informed consent was not required for this type of investigation. The study was conducted in accordance with the ethical standards laid out in the Declaration of Helsinki, 1964.

## Results

In total, among the reviewed 658 Japanese CN participants from 10 different groups (5 from original data and 5 from earlier literatures^24-28^), 103 (15.7%) turned out to be amyloid positive. The overall frequency of preclinical AD (across all stages) was estimated as 17.2% (95% CI: 12.0 – 23.1%) (forest plot in Figure 2A), with a relatively-high statistical heterogeneity of I^2^ = 64.3% (> 50%). There was no statistically-significant publication bias (*p = 0.064*).

**Figure 2.**
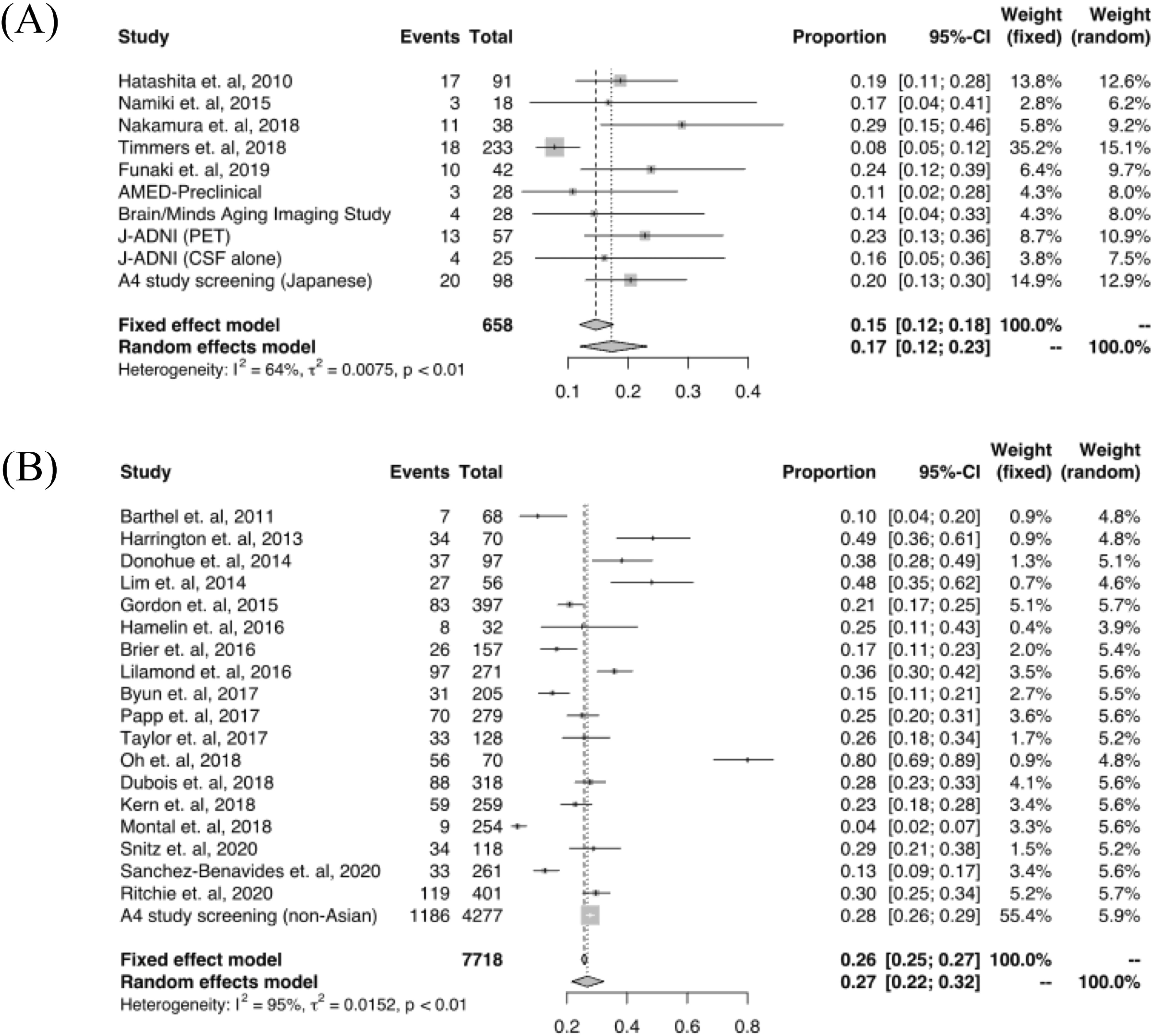
Forest plot collecting Japanese (A) and non-Japanese (B) cohorts. Forest plots meta-analyzing (A) Japanese cohorts and (B) non-Japanese cohorts, as summarized in Table 1.

And Figure 2B is the forest plot meta-analyzing non-Japanese cohorts, by additionally incorporating 18 earlier literatures data^3, 12, 29-44^ (including references in Supplemental) and one original data (of A4 screening cohort) from Western country cohorts: among the reviewed 7,718 CN participants from 21 different groups, 2,037 (26.4%) turned out to be amyloid positive. In this time, the estimated frequency of preclinical AD (of any stage) was 26.8% (95% CI: 21.7 – 32.2%) in random-effect model, with the much higher heterogeneity of I^2^ = 94.7%. There was no statistically-significant publication bias (*p = 0.923*).

Figure 3 shows a plot of mean age versus frequency of preclinical AD across all the included groups, showing a clear increase of frequency as the mean age elevates (+1.5% per-1 increase in the mean age, with *p < 0.0001* in meta-regression across all included cohorts: n = 28). In visual inspection, Japanese cohorts may look like to have a slightly lower frequency of preclinical AD than the Western cohorts have, and there was a significant difference in the frequency between Japanese and non-Japanese cohorts (*p = 0.024* in unpaired t-test) while there was no difference in the mean age (*p = 0.747* in t-test) or in the cohort sample size (*p = 0.135* in t-test). Accordingly, in meta-regression across all cohorts, being the Japanese cohort had significant association with the lower frequency of preclinical AD (estimate -8.8% with *p = 0.048* when adjusted with the mean age).

**Figure 3.**
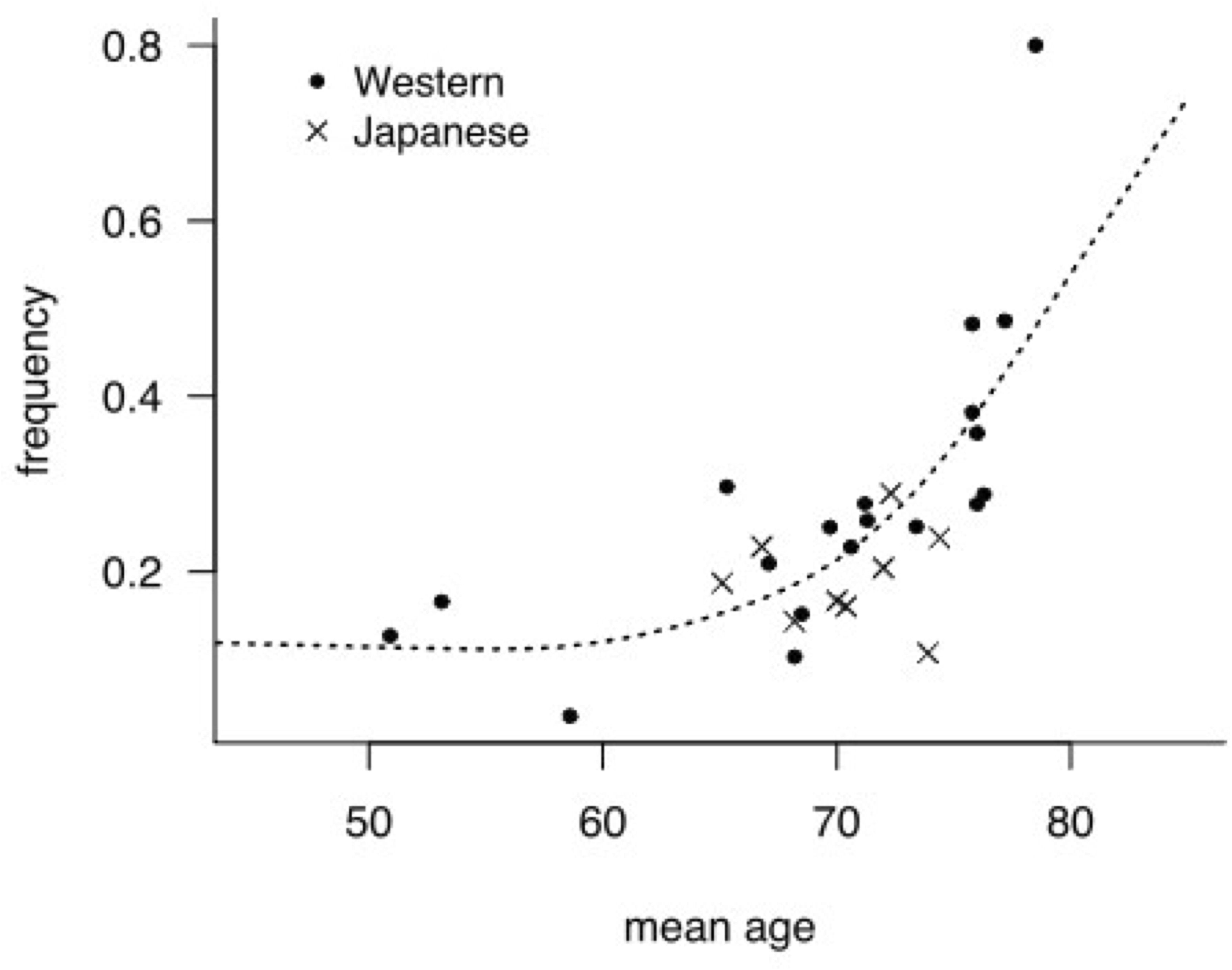
Mean age versus frequency of preclinical AD in each cohort. A plot of mean age versus frequency of preclinical AD across all the included groups. In visual inspection, Japanese cohorts may look like to have a slightly lower frequency of preclinical AD than the Western cohorts have. Spline is drawn with a dotted line. Abbreviations: AD, Alzheimer’s disease

## Discussion

In this meta-analysis, we assessed whether the frequency of preclinical AD in the clinical study settings in Japan is lower than that in non-Japanese, Western countries. This question was derived from our previous observation where preclinical AD was 23% among CN participants in J-ADNI cohort, which was lower than that in similar cohorts from Western countries (e.g., ADNI or AIBL). As a result, the frequency of preclinical AD showed clear positive correlation with the mean age, in a way that the frequency increases exponentially at 70 or older age (Figure 3): this corresponds well with the earlier meta-analysis^5^. In meta-regression, being the Japanese cohort had barely significant association with the lower frequency of preclinical AD (*p = 0.048*), of which coefficient was minus (< 0). Considering the barely low p-value and the risk of familywise error, from these results we cannot conclude that the frequency of preclinical AD in clinical study settings of Japan may be lower than that of Western countries. Our results suggest that the similar level of number of amyloid test screening might be required to acquire the same number of eligible participants for preclinical AD clinical trials in Japan as in North America or Europe, as long as we pay attention to the actual age distribution of participants.

The lower estimated prevalence of *APOE*-e4 in Japan than in North America or Central/Northern Europe^10^ may have caused the barely lower frequency of preclinical AD in Japanese cohorts, and similarly-lower frequency of preclinical AD may also be observed in other Asian countries like China or South Korea where the similarly-lower prevalence of *APOE*-e4 is estimated^10^. We would be able to validate these points during the amyloid screening process of some multi-national clinical studies/trials, such as AHEAD trial (NCT04468659) where BAN2401 (lecanemab) is to be administered to preclinical AD participants in North America, Japan, Singapore, and Europe^45^. Meanwhile, as we could not include the frequency of *APOE*-e4 genotype into the meta-regression, the lower prevalence of *APOE*-e4 in Japanese population may not be sufficient to explain the current findings: there may be some confounding factors that bring selection bias preferentially recruiting the participants with lower risk of amyloid accumulation, such as negative family history of dementia or AD^14^.

Because of the varying inclusion criteria and the varying definition of CN individuals depending on each of original cohort studies, the obtained study results cannot be interpreted as the prevalence in general population in Japan. Meanwhile, we can still interpret the synthesized results as the estimated frequency of preclinical AD (of any stage) in clinical study settings, by choosing studies of which inclusion criteria includes CDR = 0 as a minimum requirement. The CDR is one of the most critical metrics to discriminate the cognitive statuses between CN and MCI (and AD dementia)^12^. The varying inclusion criteria for each cohort study on cognitive status can then be interpreted as the minor variability of cognitive decline within the range of “cognitive normal” (CDR = 0).

Our current approach has some limitations. First, some potential confounding factors such as the frequency of *APOE*-e4 genotype or the preclinical AD staging classification are not considered due to the limited availability in earlier literatures. Second, the sample size of each cohort varied greatly (range 18-4277), leading to a large statistical heterogeneity. Third, due to the limited number of available cohorts, we collectively synthesized the cohorts with different types of amyloid test (i.e., PET and CSF) into together, which may have increased the risk of bias. Fourth, since this is not a systematic review in its strict sense, not all the eligible literatures may have been covered in the literature review. And fifth, the meta-regression results were not such robust with a barely significant p-value (*p = 0.048*), so that the identified association between the Japanese cohort and the frequency of preclinical AD may turn out to be insignificant along with the further accumulation of preclinical AD studies.

To conclude, in this study, we showed that being the Japanese cohort had a barely-significant association with the lower frequency of preclinical AD (of any stage: 1-3), suggesting that there is no robust evidence to support the lower frequency of preclinical AD in clinical study settings of Japan than that of Western countries.

## Data Availability

Original data used in this study is available from NDB or LONI databases.

https://humandbs.biosciencedbc.jp/en/hum0043-v1

https://humandbs.biosciencedbc.jp/en/hum0235-v1

https://humandbs.biosciencedbc.jp/en/hum0250-v1

https://ida.loni.usc.edu/

## Funding

This study was supported by Japan Agency for Medical Research and Development grants JP17DK0207028 and JP19DK0207048, and by the Japan Society for the Promotion of Science (JSPS) KAKENHI Grant Numbers 20J11009.

## Conflicts of interest

The authors have no conflict of interest to disclose.

## Notes

### Competing Interest Statement

The authors have declared no competing interest.

### Author Declarations

This study has been approved by the University of Tokyo Graduate School of Medicine institutional ethics committee (ID: 11628-(3)).

## References

1. Sperling RA, Aisen PS, Beckett LA, Bennett DA, Craft S, Fagan AM, Iwatsubo T, Jack CR Jr, Kaye J, Montine TJ, Park DC, Reiman EM, Rowe CC, Siemers E, Stern Y, Yaffe K, Carrillo MC, Thies B, Morrison-Bogorad M, Wagster MV, Phelps CH. Toward defining the preclinical stages of Alzheimer’s disease: recommendations from the National Institute on Aging-Alzheimer’s Association workgroups on diagnostic guidelines for Alzheimer’s disease. Alzheimers Dement. 2011 May;7(3):280–92. doi: 10.1016/j.jalz.2011.03.003. Epub 2011 Apr 21. PMID: 21514248; PMCID: PMC3220946.

2. Jack CR Jr, Knopman DS, Jagust WJ, Petersen RC, Weiner MW, Aisen PS, Shaw LM, Vemuri P, Wiste HJ, Weigand SD, Lesnick TG, Pankratz VS, Donohue MC, Trojanowski JQ. Tracking pathophysiological processes in Alzheimer’s disease: an updated hypothetical model of dynamic biomarkers. Lancet Neurol. 2013 Feb;12(2):207–16. doi: 10.1016/S1474-4422(12)70291-0. PMID: 23332364; PMCID: PMC3622225.

3. Donohue MC, Sperling RA, Salmon DP, Rentz DM, Raman R, Thomas RG, Weiner M, Aisen PS; Australian Imaging, Biomarkers, and Lifestyle Flagship Study of Ageing; Alzheimer’s Disease Neuroimaging Initiative; Alzheimer’s Disease Cooperative Study. The preclinical Alzheimer cognitive composite: measuring amyloid-related decline. JAMA Neurol. 2014 Aug;71(8):961–70. doi: 10.1001/jamaneurol.2014.803. PMID: 24886908; PMCID: PMC4439182.

4. Cummings J, Lee G, Ritter A, Sabbagh M, Zhong K. Alzheimer’s disease drug development pipeline: 2020. Alzheimers Dement (N Y). 2020 Jul 16;6(1):e12050. doi: 10.1002/trc2.12050. PMID: 32695874; PMCID: PMC7364858.

5. Jansen WJ, Ossenkoppele R, Knol DL, Tijms BM, Scheltens P, Verhey Fr, Visser PJ; Amyloid Biomarker Study Group, Aalten P, Aarsland D, Alcolea D, Alexander M, Almdahl IS, Arnold SE, Baldeiras I, Barthel H, van Berckel BN, Bibeau K, Blennow K, Brooks DJ, van Buchem MA, Camus V, Cavedo E, Chen K, Chetelat G, Cohen AD, Drzezga A, Engelborghs S, Fagan AM, Fladby T, Fleisher AS, van der Flier WM, Ford L, Förster S, Fortea J, Foskett N, Frederiksen KS, Freund-Levi Y, Frisoni GB, Froelich L, Gabryelewicz T, Gill KD, Gkatzima O, Gómez-Tortosa E, Gordon MF, Grimmer T, Hampel H, Hausner L, Hellwig S, Herukka SK, Hildebrandt H, Ishihara L, Ivanoiu A, Jagust WJ, Johannsen P, Kandimalla R, Kapaki E, Klimkowicz-Mrowiec A, Klunk WE, Köhler S, Koglin N, Kornhuber J, Kramberger MG, Van Laere K, Landau SM, Lee DY, de Leon M, Lisetti V, Lleó A, Madsen K, Maier W, Marcusson J, Mattsson N, de Mendonça A, Meulenbroek O, Meyer PT, Mintun MA, Mok V, Molinuevo JL, Møllergård HM, Morris JC, Mroczko B, Van der Mussele S, Na DL, Newberg A, Nordberg A, Nordlund A, Novak GP, Paraskevas GP, Parnetti L, Perera G, Peters O, Popp J, Prabhakar S, Rabinovici GD, Ramakers IH, Rami L, Resende de Oliveira C, Rinne JO, Rodrigue KM, Rodríguez-Rodríguez E, Roe CM, Rot U, Rowe CC, Rüther E, Sabri O, Sanchez-Juan P, Santana I, Sarazin M, Schröder J, Schütte C, Seo SW, Soetewey F, Soininen H, Spiru L, Struyfs H, Teunissen CE, Tsolaki M, Vandenberghe R, Verbeek MM, Villemagne VL, Vos SJ, van Waalwijk van Doorn LJ, Waldemar G, Wallin A, Wallin ÅK, Wiltfang J, Wolk DA, Zboch M, Zetterberg H. Prevalence of cerebral amyloid pathology in persons without dementia: a meta-analysis. JAMA. 2015 May 19;313(19):1924–38. doi: 10.1001/jama.2015.4668. PMID: 25988462; PMCID: PMC4486209.

6. Rowe CC, Ellis KA, Rimajova M, Bourgeat P, Pike KE, Jones G, Fripp J, Tochon-Danguy H, Morandeau L, O’Keefe G, Price R, Raniga P, Robins P, Acosta O, Lenzo N, Szoeke C, Salvado O, Head R, Martins R, Masters CL, Ames D, Villemagne VL. Amyloid imaging results from the Australian Imaging, Biomarkers and Lifestyle (AIBL) study of aging. Neurobiol Aging. 2010 Aug;31(8):1275–83. doi: 10.1016/j.neurobiolaging.2010.04.007. Epub 2010 May 15. PMID: 20472326.

7. Parnetti L, Chipi E, Salvadori N, D’Andrea K, Eusebi P. Prevalence and risk of progression of preclinical Alzheimer’s disease stages: a systematic review and meta-analysis. Alzheimers Res Ther. 2019 Jan 15;11(1):7. doi: 10.1186/s13195-018-0459-7. PMID: 30646955; PMCID: PMC6334406.

8. Iwatsubo T, Iwata A, Suzuki K, Ihara R, Arai H, Ishii K, Senda M, Ito K, Ikeuchi T, Kuwano R, Matsuda H; Japanese Alzheimer’s Disease Neuroimaging Initiative, Sun CK, Beckett LA, Petersen RC, Weiner MW, Aisen PS, Donohue MC; Alzheimer’s Disease Neuroimaging Initiative. Japanese and North American Alzheimer’s Disease Neuroimaging Initiative studies: Harmonization for international trials. Alzheimers Dement. 2018 Aug;14(8):1077–1087. doi: 10.1016/j.jalz.2018.03.009. Epub 2018 May 9. PMID: 29753531.

9. Ihara R, Iwata A, Suzuki K, Ikeuchi T, Kuwano R, Iwatsubo T; Japanese Alzheimer’s Disease Neuroimaging Initiative. Clinical and cognitive characteristics of preclinical Alzheimer’s disease in the Japanese Alzheimer’s Disease Neuroimaging Initiative cohort. Alzheimers Dement (N Y). 2018 Nov 26;4:645–651. doi: 10.1016/j.trci.2018.10.004. PMID: 30511010; PMCID: PMC6258138.

10. Ward A, Crean S, Mercaldi CJ, Collins JM, Boyd D, Cook MN, Arrighi HM. Prevalence of apolipoprotein E4 genotype and homozygotes (APOE e4/4) among patients diagnosed with Alzheimer’s disease: a systematic review and meta-analysis. Neuroepidemiology. 2012;38(1):1–17. doi: 10.1159/000334607. Epub 2011 Dec 17. PMID: 22179327.

11. Jack CR Jr, Knopman DS, Weigand SD, Wiste HJ, Vemuri P, Lowe V, Kantarci K, Gunter JL, Senjem ML, Ivnik RJ, Roberts RO, Rocca WA, Boeve BF, Petersen RC. An operational approach to National Institute on Aging-Alzheimer’s Association criteria for preclinical Alzheimer disease. Ann Neurol. 2012 Jun;71(6):765–75. doi: 10.1002/ana.22628. Epub 2012 Apr 9. PMID: 22488240; PMCID: PMC3586223.

12. Kern S, Zetterberg H, Kern J, Zettergren A, Waern M, Höglund K, Andreasson U, Wetterberg H, Börjesson-Hanson A, Blennow K, Skoog I. Prevalence of preclinical Alzheimer disease: Comparison of current classification systems. Neurology. 2018 May 8;90(19):e1682–e1691. doi: 10.1212/WNL.0000000000005476. Epub 2018 Apr 13. PMID: 29653987; PMCID: PMC5952969.

13. Sperling RA, Rentz DM, Johnson KA, Karlawish J, Donohue M, Salmon DP, Aisen P. The A4 study: stopping AD before symptoms begin? Sci Transl Med. 2014 Mar 19;6(228):228fs13. doi: 10.1126/scitranslmed.3007941. PMID: 24648338; PMCID: PMC4049292.

14. Sperling RA, Donohue MC, Raman R, Sun CK, Yaari R, Holdridge K, Siemers E, Johnson KA, Aisen PS; A4 Study Team. Association of Factors With Elevated Amyloid Burden in Clinically Normal Older Individuals. JAMA Neurol. 2020 Jun 1;77(6):735–745. doi: 10.1001/jamaneurol.2020.0387. PMID: 32250387; PMCID: PMC7136861.

15. Petersen RC, Aisen PS, Beckett LA, Donohue MC, Gamst AC, Harvey DJ, Jack CR Jr, Jagust WJ, Shaw LM, Toga AW, Trojanowski JQ, Weiner MW. Alzheimer’s Disease Neuroimaging Initiative (ADNI): clinical characterization. Neurology. 2010 Jan 19;74(3):201–9. doi: 10.1212/WNL.0b013e3181cb3e25. Epub 2009 Dec 30. PMID: 20042704; PMCID: PMC2809036.

16. Iwata A, Iwatsubo T, Ihara R, Suzuki K, Matsuyama Y, Tomita N, Arai H, Ishii K, Senda M, Ito K, Ikeuchi T, Kuwano R, Matsuda H; Alzheimer’s Disease Neuroimaging Initiative; Japanese Alzheimer’s Disease Neuroimaging Initiative. Effects of sex, educational background, and chronic kidney disease grading on longitudinal cognitive and functional decline in patients in the Japanese Alzheimer’s Disease Neuroimaging Initiative study. Alzheimers Dement (N Y). 2018 Jul 12;4:765–774. doi: 10.1016/j.trci.2018.06.008. PMID: 30662934; PMCID: PMC6324255.

17. Sato K, Mano T, Ihara R, Suzuki K, Tomita N, Arai H, Ishii K, Senda M, Ito K, Ikeuchi T, Kuwano R, Matsuda H, Iwatsubo T, Toda T, Iwata A; Alzheimer’s Disease Neuroimaging Initiative, and Japanese Alzheimer’s Disease Neuroimaging Initiative. Lower Serum Calcium as a Potentially Associated Factor for Conversion of Mild Cognitive Impairment to Early Alzheimer’s Disease in the Japanese Alzheimer’s Disease Neuroimaging Initiative. J Alzheimers Dis. 2019;68(2):777–788. doi: 10.3233/JAD-181115. PMID: 30814351.

18. Sato K, Mano T, Matsuda H, Senda M, Ihara R, Suzuki K, Arai H, Ishii K, Ito K, Ikeuchi T, Kuwano R, Toda T, Iwatsubo T, Iwata A; Japanese Alzheimer’s Disease Neuroimaging Initiative. Visualizing modules of coordinated structural brain atrophy during the course of conversion to Alzheimer’s disease by applying methodology from gene co-expression analysis. Neuroimage Clin. 2019;24:101957. doi: 10.1016/j.nicl.2019.101957. Epub 2019 Jul 25. PMID: 31400633; PMCID: PMC6700430.

19. Kenichiro Sato, Ryoko Ihara, Kazushi Suzuki, Yoshiki Niimi, Tatsushi Toda, Gustavo Jimenez-Maggiora, Oliver Langford, Michael C. Donohue, Rema Raman, Paul S. Aisen, Reisa A. Sperling, Atsushi Iwata, Takeshi Iwatsubo. Predicting amyloid risk by machine learning algorithms based on the A4 screen data: application to the Japanese Trial-Ready Cohort study. Alzheimer’s & Dementia: Translational Research & Clinical Interventions. 2021. doi:10.1002/trc2.12135

20. Balduzzi S, Rücker G, Schwarzer G (2019), How to perform a meta-analysis with R: a practical tutorial, Evidence-Based Mental Health; 22: 153–160.

21. Viechtbauer, W. (2010). Conducting meta-analyses in R with the metafor package. Journal of Statistical Software, 36(3), 1-48. URL: https://www.jstatsoft.org/v36/i03/

22. Egger M, Davey Smith G, Schneider M, Minder C. Bias in meta-analysis detected by a simple, graphical test. BMJ. 1997 Sep 13;315(7109):629–34. doi: 10.1136/bmj.315.7109.629. PMID: 9310563; PMCID: PMC2127453.

23. Wood, S.N. (2017) Generalized Additive Models: An Introduction with R (2nd edition). Chapman and Hall/CRC.

24. Hatashita S, Yamasaki H. Clinically different stages of Alzheimer’s disease associated by amyloid deposition with [11C]-PIB PET imaging. J Alzheimers Dis. 2010;21(3):995–1003. doi: 10.3233/JAD-2010-100222. PMID: 20693641.

25. Namiki C, Takita Y, Iwata A, Momose T, Senda M, Okubo Y, Joshi AD, Lu M, Agbulos A, Breault C, Pontecorvo MJ. Imaging characteristics and safety of florbetapir (^1^□F) in Japanese healthy volunteers, patients with mild cognitive impairment and patients with Alzheimer’s disease. Ann Nucl Med. 2015 Aug;29(7):570–81. doi: 10.1007/s12149-015-0978-2. Epub 2015 May 6. PMID: 25943346.

26. Nakamura A, Cuesta P, Fernández A, Arahata Y, Iwata K, Kuratsubo I, Bundo M, Hattori H, Sakurai T, Fukuda K, Washimi Y, Endo H, Takeda A, Diers K, Bajo R, Maestú F, Ito K, Kato T. Electromagnetic signatures of the preclinical and prodromal stages of Alzheimer’s disease. Brain. 2018 May 1;141(5):1470–1485. doi: 10.1093/brain/awy044. PMID: 29522156; PMCID: PMC5920328.

27. Timmers M, Streffer JR, Russu A, Tominaga Y, Shimizu H, Shiraishi A, Tatikola K, Smekens P, Börjesson-Hanson A, Andreasen N, Matias-Guiu J, Baquero M, Boada M, Tesseur I, Tritsmans L, Van Nueten L, Engelborghs S. Pharmacodynamics of atabecestat (JNJ-54861911), an oral BACE1 inhibitor in patients with early Alzheimer’s disease: randomized, double-blind, placebo-controlled study. Alzheimers Res Ther. 2018 Aug 23;10(1):85. doi: 10.1186/s13195-018-0415-6. PMID: 30134967; PMCID: PMC6106931.

28. Funaki K, Nakajima S, Noda Y, Wake T, Ito D, Yamagata B, Yoshizaki T, Kameyama M, Nakahara T, Murakami K, Jinzaki M, Mimura M, Tabuchi H. Can we predict amyloid deposition by objective cognition and regional cerebral blood flow in patients with subjective cognitive decline? Psychogeriatrics. 2019 Jul;19(4):325–332. doi: 10.1111/psyg.12397. Epub 2019 Jan 27. PMID: 30688000.

29. Lozupone M, Solfrizzi V, D’Urso F, Di Gioia I, Sardone R, Dibello V, Stallone R, Liguori A, Ciritella C, Daniele A, Bellomo A, Seripa D, Panza F. Anti-amyloid-β protein agents for the treatment of Alzheimer’s disease: an update on emerging drugs. Expert Opin Emerg Drugs. 2020 Sep;25(3):319–335. doi: 10.1080/14728214.2020.1808621. Epub 2020 Aug 20. PMID: 32772738.

